# Unresolved Cardiometabolic Care Gaps and Cardiovascular Outcomes: A Count–Stage Framework Across 3 Prospective Cohorts

**DOI:** 10.64898/2026.11.18.26368455

**Authors:** Yunxi Yang, Jia Lv, Xinyi Tang, Xinghua Zheng, Celing Xv, Xianchang Lai, Xing Liu

## Abstract

**BACKGROUND:** People with the same number of unresolved cardiometabolic problems may be at different stages of care. Whether care stage provides information on subsequent cardiovascular risk beyond the number of unresolved problems remains unclear.

**METHODS:** We analyzed longitudinal data from CHARLS, HRS, and ELSA. Measurements, diagnoses, and treatment information were used to classify care states at each landmark. Cohort-specific adjusted Cox models estimated associations with subsequent cardiovascular disease (CVD), heart disease, and stroke. We tested whether stage added information beyond gap count and whether count remained informative after stage was considered. We also directly compared blood pressure, glycemic, and lipid targets within the same stage.

**RESULTS:** CHARLS included 7,899 participants. HRS and ELSA included 20,531 and 12,071 landmark observations from 12,933 and 6,491 participants, respectively. Hazard ratios (HRs) and 95% CIs for CVD per additional gap were 1.31 (1.23–1.41), 1.18 (1.13–1.23), and 1.09 (1.03–1.16). With exactly 1 gap, HRs for treated but uncontrolled (TA) versus unrecognized (U) gaps were 2.09 (1.70–2.57), 1.34 (1.19–1.51), and 1.27 (1.07–1.50) after adjustment for the affected target. Single-gap care stage provided additional information on CVD and heart disease risk beyond gap count in all 3 cohorts. Outcomes also differed between 2 and 3 gaps after accounting for stage in CHARLS and HRS. Under cohort-specific definitions, TA was the most common glycemic gap stage in all 3 cohorts, whereas U predominated among lipid gaps in CHARLS and ELSA. CVD associations were stronger for unrecognized glycemic gaps than for unrecognized blood pressure or lipid gaps in CHARLS. At the TA stage in HRS, associations were stronger for blood pressure and glycemic gaps than for lipid gaps.

**CONCLUSIONS:** Gap count and care stage provide complementary prognostic information. The count–stage framework first records how many cardiometabolic targets remain unresolved, then identifies each affected target and its care stage to describe where further assessment is needed.

**CLINICAL PERSPECTIVE:** *What Is New?:*

- In CHARLS, HRS, and ELSA, gap count and care stage provided complementary prognostic information on unresolved cardiometabolic problems. With exactly 1 gap, treated but uncontrolled (TA) gaps were associated with higher CVD risk than unrecognized (U) gaps, even after adjustment for the affected target.
- For multiple gaps, grouping combinations as same-stage or mixed-stage did not consistently distinguish subsequent outcomes. Among observations with exactly 2 gaps, combinations containing TA were associated with higher CVD risk in each cohort.

*What Are the Clinical Implications?:*

- Gap count records how many targets remain unresolved. Care stage helps identify whether further assessment concerns diagnostic confirmation, treatment initiation, or control during treatment.
- A concise description of multiple care gaps can retain the exact count, affected targets, and presence of TA. Implementation studies could evaluate whether this approach helps clinicians organize follow-up.

## 1 Introduction

Blood pressure, glycemic, and lipid abnormalities often coexist in middle-aged and older adults and are major targets for long-term cardiovascular prevention.^1–4^ The same abnormal measurement can call for different follow-up assessments. A person with elevated blood pressure who has never been diagnosed with hypertension needs diagnostic confirmation. Someone whose blood pressure remains elevated during antihypertensive treatment needs an assessment of why control has not been achieved.^5–8^ Clinicians therefore need to know how many targets remain unresolved and whether each problem concerns recognition, treatment initiation, or control during treatment.

Care cascade studies have described recognition, treatment, and control, including the joint management of several cardiovascular risk factors.^9–13^ Lawton and Cutler considered both population risk factor burden and care among people at risk, showing how each contributes to the proportion with uncontrolled risk.^9^ Studies of hypertension have also linked care stages to subsequent cardiovascular outcomes.^14^ Research on combined risk factors and metabolic indices has largely focused on the risks associated with accumulating abnormalities.^15–19^ Whether care stage distinguishes subsequent risk among people with the same number of unresolved problems remains less clear. It is also uncertain whether blood pressure, glycemic, and lipid gaps have similar associations with outcomes when they occur at the same stage of care.

Disease history does not fully describe current care needs. A diagnosed and controlled condition still requires follow-up, but it does not constitute a current care gap. An undiagnosed abnormality may represent a care gap despite being absent from the recorded disease history. Counting diagnoses therefore misses unrecognized abnormalities, while measurements alone do not show whether the next assessment concerns recognition, treatment initiation, or control during treatment. When several targets remain unresolved, the description becomes more complex. It remains unclear whether distinguishing stage combinations adds useful information at a fixed gap count.

We used longitudinal data from CHARLS, HRS, and ELSA to examine unresolved blood pressure, glycemic, and lipid problems as care gaps at each landmark. We assessed their associations with subsequent CVD, heart disease, and stroke, asking whether stage added information beyond exact gap count and whether count retained information after stage was considered. We also compared targets within the same care stage and examined whether stage combinations were associated with different outcomes at a fixed count among observations with multiple gaps. These analyses informed a count–stage framework that first records how many targets remain unresolved and then describes the care stage of each affected target.

## 2 Methods

### 2.1 Data Sources, Study Design, and Population

The China Health and Retirement Longitudinal Study (CHARLS), Health and Retirement Study (HRS), and English Longitudinal Study of Ageing (ELSA) are nationally representative longitudinal studies of middle-aged and older adults in China, the United States, and England, respectively.^20–22^ HRS began in 1992 and conducts follow-up approximately every 2 years. ELSA has followed adults aged 50 years or older in England approximately every 2 years since 2002 using a design harmonized with HRS. CHARLS has followed Chinese adults aged 45 years or older since 2011, generally at intervals of 2 to 3 years. All collect repeated information on demographic characteristics, disease diagnoses, medication use, and health measurements. We applied a common conceptual care framework using the measurements, diagnosis, and treatment information available in each cohort. Ethics approval and informed consent for the original studies are described at the end of this article.

Measurement visits served as landmarks. Exposures and covariates were assessed at each visit, and follow-up began thereafter.^20–22^ In CHARLS, eligible participants were aged ≥45 years in 2015, were confirmed free of heart disease and stroke at that visit, and had no CVD report at any available earlier visit. Care gaps were assessed in 2015, with earlier visits used to establish prior CVD history. Participation in 2011 and complete gap classification in that year were not required. HRS contributed biomarker landmarks from 2006–2016 among adults aged ≥50 years, and ELSA contributed nurse assessment landmarks at Waves 2, 4, 6, and 8 among adults aged ≥50 years. In all cohorts, eligibility required absence of CVD at the landmark, classifiable states for all 3 targets, complete covariates for the fitted model, and valid follow-up for the relevant outcome (Figure 1 and Tables S7A–F). HRS and ELSA participants could contribute multiple landmarks, each opening a new follow-up window. The same first-reported event could therefore contribute to more than 1 overlapping landmark follow-up window. Models accounted for clustering by participant, and we report both landmark observations and unique participants. Reporting followed the STROBE guidelines.^23^

**Figure 1.**
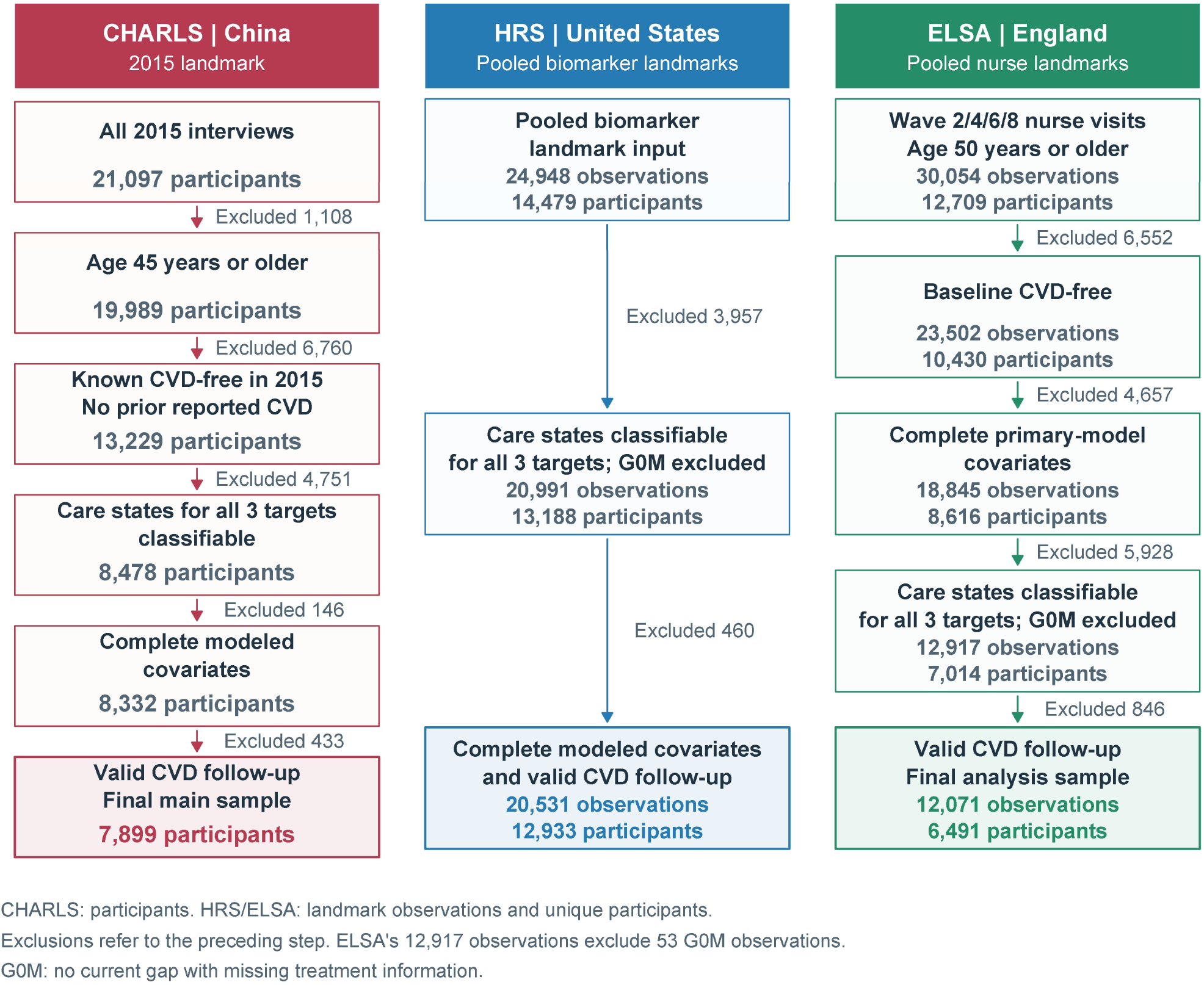
Selection of the study population in the 3 cohorts. CHARLS reports participants; HRS and ELSA report landmark observations and unique participants; exclusions are calculated relative to the preceding step. G0M denotes no current gap with missing treatment information, preventing classification as U0 or TC. The 12,917 ELSA observations already exclude 53 G0M observations. Selection sensitivity analyses are reported in Tables S7G1–G2 and S7H1–H2.

### 2.2 Gap Count and Care Stage

We combined current measurements with prior diagnosis and treatment information to assign each target to 1 of 5 care states. Targets with no current gap were classified as untreated (U0) or treated and currently controlled (TC). Current care gaps were classified as unrecognized (U), recognized but untreated (RU), or treated but uncontrolled (TA), according to cohort-specific measurement, diagnosis, and treatment criteria. These 3 gap categories described the care stage of each unresolved target. All cohorts defined a blood pressure gap as systolic blood pressure ≥140 mm Hg or diastolic blood pressure ≥90 mm Hg. In CHARLS, glycemic classification gave priority to current control, defined by HbA1c <7.0%. Participants with an available HbA1c below this threshold were classified as having no current glycemic gap, including those with HbA1c from 6.5% to <7.0%. Fasting glucose ≥126 mg/dL was used to identify lack of control only when HbA1c was missing. HRS used HbA1c ≥6.5%; ELSA used the same threshold, with fasting glucose ≥7.0 mmol/L as a substitute when HbA1c was missing. Tables S1A1–S1B give the cohort-specific indicators, order of classification, and missing-data rules. In CHARLS, participants classified as having a current gap were assigned to U if they were untreated and had no reported diagnosis, including those with missing diagnosis responses. In HRS, observations with a gap and missing treatment responses were assigned to U or RU according to known diagnosis status. We examined the effect of excluding these observations in Table S7H1.

Summing gaps across the 3 targets gave a count of 0, 1, 2, or 3. We also defined the overall phenotypes G0, U, RU, TA, and M. Overall G0 required all targets to be free of a current gap, with each classified as U0 or TC. Overall U, RU, and TA represented exactly 1 gap at the corresponding stage. M included every stage combination with 2–3 gaps. For analyses of an individual target, G0 combined U0 and TC for that target. Care stages were modeled as separate nominal categories.

### 2.3 Outcomes and Primary Models

The primary outcome was the first reported CVD after the landmark, defined as heart disease or stroke. These 2 conditions were also analyzed separately as secondary outcomes. Heart disease was identified using each cohort’s composite questionnaire measure. Event time was approximated by the survey at which an outcome was first reported, and observations without an event were censored at the end of valid follow-up for that outcome. CHARLS follow-up included the 2018 and 2020 surveys, 3 and 5 years after the landmark. HRS follow-up extended to 2022 and ELSA follow-up to Wave 11. Questionnaire items, outcome construction, and censoring rules are described in Supplemental Material S1. Table S2A reports denominators, event counts, and follow-up summaries.

We estimated hazard ratios (HRs) and 95% confidence intervals (CIs) using adjusted Cox models fitted separately in each cohort. All models included age, sex, education, marital status, smoking, alcohol use, and body mass index (BMI). CHARLS models additionally included urban–rural residence and kidney disease, HRS models included region and landmark year, and ELSA models included survey wave (Table S1C). We used model-based standard errors in CHARLS and participant-clustered robust standard errors in HRS and ELSA. Gap count was entered as categories of 0, 1, 2, and 3, using 0 as the reference, and as a continuous integer to estimate the association per additional gap. Overall phenotype models used overall G0 as the reference. Target-specific 5-state models used U0 as the reference and included TC. We used unweighted complete-case analyses to estimate associations within each cohort. Missing exposure, covariate, and outcome-related data were not multiply imputed. Reported proportions describe the included observations. Figure 1 and Tables S7A–F show the selection process and comparisons of included and excluded observations.

### 2.4 Stage Information and Comparisons Across Targets

To test whether stage added information beyond count, we extended the exact-count model by distinguishing U, RU, and TA among observations with exactly 1 gap. To test whether count added information beyond stage classification, we extended the overall 5-category model by separating M into 2 and 3 gaps. Joint Wald tests used the variance estimator of the primary models. Holm correction was applied across the 3 outcomes within each cohort and comparison. We evaluated gains in model fit with 500 participant-level bootstrap resamples and report apparent ΔC-index values calculated in the same sample (Supplemental Material S1 and Tables S3B1–S3B2).

For observations with exactly 1 gap, we compared RU and TA with U using the primary-model covariates and then additionally adjusted for the affected target, classified as blood pressure, glycemia, or lipids. For multiple gaps, we compared mixed-stage with same-stage combinations and combinations containing at least 1 TA gap with those containing none. Models restricted to exactly 2 gaps adjusted for target composition. Models including observations with 2–3 gaps adjusted for exact gap count (Tables S3A2–S3C2).

Direct comparisons among targets within the same stage used each target’s G0 as the reference. We fitted stacked models stratified by target, allowing both stage and covariate coefficients to vary by target, with covariance estimates clustered by participant. Overall Wald tests and pairwise contrasts assessed differences among targets. To examine the reference group, we separated target-specific G0 into U0 and TC and compared TA with TC in 5-state models. We also omitted each target in turn and reconstructed overall phenotypes from the remaining targets. Stroke and heart disease associations were compared as ratios of HRs (RHRs) using stacked models stratified by outcome (Tables S4A–S5C).

We pooled estimates from the 3 cohorts using restricted maximum likelihood (REML) random-effects models with Hartung–Knapp inference.^24^ The pooled tables also report heterogeneity and prediction intervals. Holm correction was applied within the comparison families specified in each table. Confidence and prediction intervals were not adjusted for multiple comparisons. We assessed sensitivity to covariate adjustment using a common minimal adjustment set and to functional form using natural splines for age and BMI. Fixed-effect meta-analysis and leave-one-cohort-out analyses assessed the influence of the pooling method and individual cohorts. Selection analyses examined earlier-visit requirements in CHARLS, missing treatment responses in HRS, and a requirement for complete urban–rural residence data in HRS. Proportional hazards tests and exposure-by-log-time interactions used the same samples as the corresponding models. Analyses were performed in R 4.3.3. Software and implementation details are provided in Supplemental Material S1, S6, and S7.

## 3 Results

### 3.1 Study Population and Distribution of Care Gaps

The analysis included 7,899 participants in CHARLS, 20,531 landmark observations from 12,933 participants in HRS, and 12,071 landmark observations from 6,491 participants in ELSA (Figure 1 and Table 1). Mean ages were 60.8, 65.6, and 65.1 years, respectively. Median follow-up within CVD windows was 5.0 years in CHARLS, 8.0 years in HRS, and 8.0 years in ELSA. CVD was recorded in 1,446, 4,554, and 2,978 follow-up windows, involving 1,446, 2,961, and 1,673 unique participants, respectively. Table 1 and Tables S2A and S7E give outcome-specific denominators, follow-up, and event-window counts.

**Table 1.** Primary Analysis Samples, Follow-Up, and Outcome Counts Across the 3 Cohorts.

| **Characteristic** | **CHARLS**<br>**China** | **HRS**<br>**United States** | **ELSA**<br>**England** |
|---|---|---|---|
| Primary analysis sample | 7,899 participants | 20,531 landmark observations | 12,071 landmark observations |
| Unique participants | 7,899 | 12,933 | 6,491 |
| CVD follow-up, person-years | 36,753 | 159,254 | 110,700 |
| Follow-up per CVD window, y, median (IQR) | 5.0 (5.0–5.0) | 8.0 (6.0–10.0) | 8.0 (6.0–12.0) |
| CVD event observations | 1,446 | 4,554 | 2,978 |
| Unique participants with CVD | 1,446 | 2,961 | 1,673 |
| Heart disease event observations | 1,055 | 3,801 | 2,619 |
| Unique participants with heart disease | 1,055 | 2,456 | 1,463 |
| Stroke event observations | 510 | 1,238 | 613 |
| Unique participants with stroke | 510 | 840 | 370 |
| Age, y, mean (SD) | 60.8 (9.0) | 65.6 (9.9) | 65.1 (8.5) |
| Female, n (%) | 4,177 (52.9) | 12,499 (60.9) | 6,524 (54.0) |
| BMI, kg/m², mean (SD) | 23.7 (3.6) | 28.7 (5.9) | 27.9 (4.9) |
| Education (cohort-specific) | High school or above, 811 (10.3%) | Years of schooling, 12.9 (3.1) | College or above, 5,434 (45.0%) |
| Married/partnered, n (%) | 6,934 (87.8) | 12,365 (60.2) | 9,072 (75.2) |
| Ever smoker, n (%) | 3,423 (43.3) | 10,944 (53.3) | 7,130 (59.1) |
| Alcohol use, n (%) | 3,666 (46.4) | 12,213 (59.5) | 8,581 (71.1) |
| 0 current gaps, n (%) | 2,967 (37.6) | 10,088 (49.1) | 3,302 (27.4) |
| 1 current gap, n (%) | 3,601 (45.6) | 7,835 (38.2) | 5,842 (48.4) |
| 2 current gaps, n (%) | 1,185 (15.0) | 2,335 (11.4) | 2,710 (22.5) |
| 3 current gaps, n (%) | 146 (1.8) | 273 (1.3) | 217 (1.8) |
| At least 1 current gap, n (%) | 4,932 (62.4) | 10,443 (50.9) | 8,769 (72.6) |

CHARLS contributed 1 landmark observation per participant. HRS and ELSA counts refer to landmark observations with follow-up windows that may overlap. Event observations and unique participants with events are reported separately. Person-years and median follow-up refer to CVD windows. The HRS stroke denominator is 20,530. Continuous variables are reported as mean (SD) and categorical variables as n (%); education retains cohort-specific definitions. Complete stratified baseline characteristics and selection data are provided in Tables S2B1–S2B3 and S7A–D.

At least 1 care gap was present in 62.4% of CHARLS participants, 50.9% of HRS landmark observations, and 72.6% of ELSA landmark observations. Approximately two-thirds to three-quarters of observations with a gap had only 1 unresolved target. Most observations with multiple gaps involved 2 targets, while 3 gaps accounted for less than 2% of all observations in each cohort. Exact counts and baseline characteristics by gap count are reported in Tables S2B1–S2B4.

The most common care stage differed across targets and cohorts. In CHARLS and ELSA, most lipid gaps were unrecognized. These U-stage lipid gaps accounted for 40.2% of CHARLS participants and 40.9% of ELSA landmark observations. U was also the most common stage for blood pressure gaps in these cohorts. In HRS, treated but uncontrolled gaps predominated for both blood pressure and lipids.

Glycemic gaps were most often at the TA stage in all 3 cohorts. Under the respective cohort definitions, TA accounted for 48.6%, 68.4%, and 59.4% of glycemic gaps in CHARLS, HRS, and ELSA. Figure S1 and Table S2D show stage distributions among target-specific gaps and the proportion of the primary analysis sample at each stage.

### 3.2 Gap Count and Cardiovascular Outcomes

A greater number of care gaps was generally associated with higher CVD risk in all 3 cohorts (Figure 2 and Table 2A). HRs per additional gap were 1.31 (1.23–1.41) in CHARLS, 1.18 (1.13–1.23) in HRS, and 1.09 (1.03–1.16) in ELSA. Compared with 0 gaps, 3 gaps were associated with CVD HRs of 2.49 (1.86–3.33) in CHARLS and 1.93 (1.54–2.40) in HRS. In ELSA, estimates were similar for 2 and 3 gaps.

**Figure 2.**
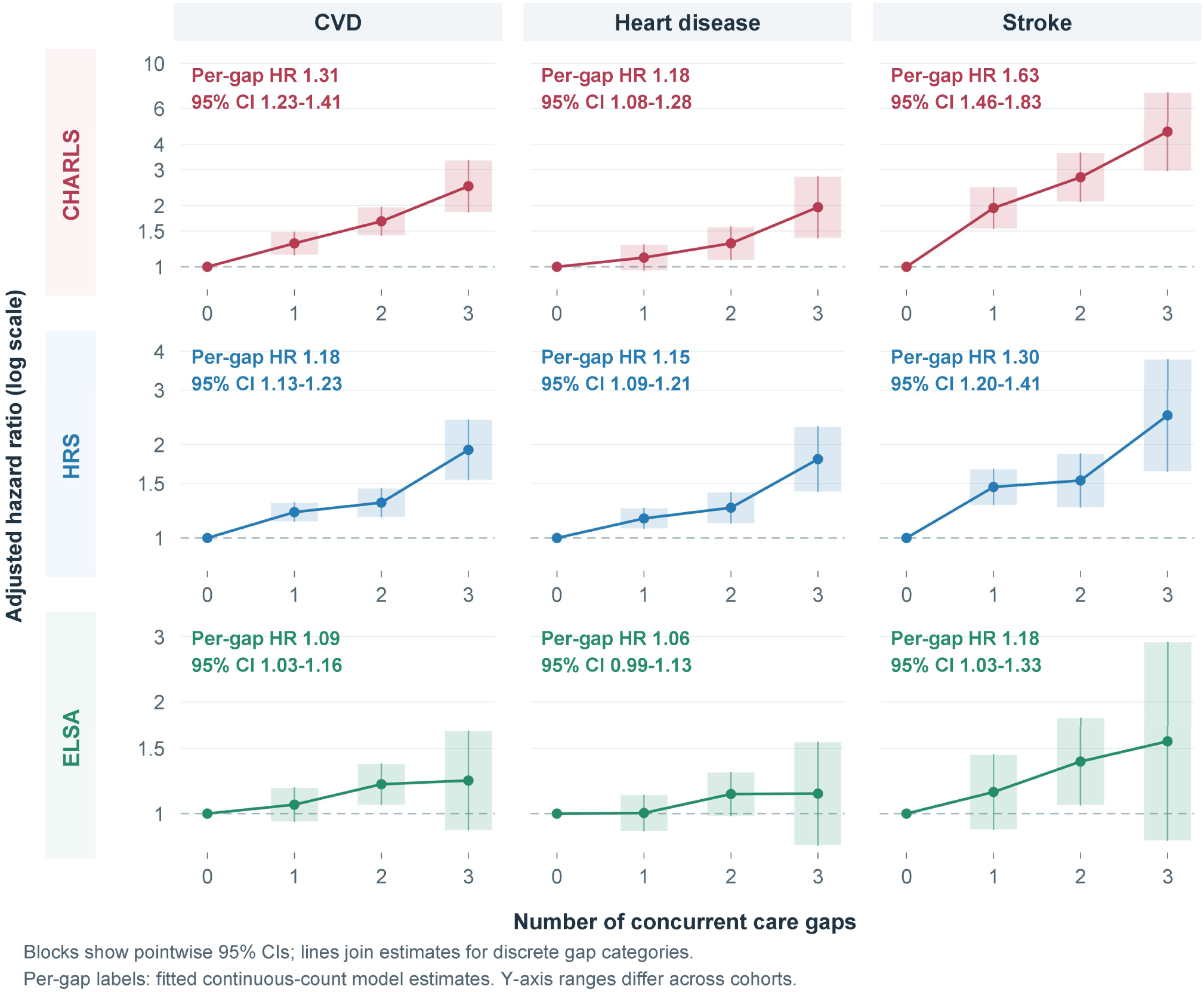
Exact gap count and cardiovascular outcomes. Zero gaps are the reference. Points and pointwise 95% CIs are from cohort-specific adjusted Cox models; lines connect the discrete categories. The vertical axis is logarithmic. Per-gap HRs are from continuous-count models, and categorical estimates are reported in Table 2A.

**Table 2A.** Adjusted Associations of Exact Gap Count With Cardiovascular Outcomes.

| **Exposure** | **Outcome** | **CHARLS HR (95% CI)** | **HRS HR (95% CI)** | **ELSA HR (95% CI)** |
|---|---|---|---|---|
| 1 gap vs 0 gaps | CVD | 1.30 (1.15–1.48) | 1.21 (1.13–1.30) | 1.06 (0.95–1.17) |
| 1 gap vs 0 gaps | Heart disease | 1.11 (0.96–1.28) | 1.16 (1.07–1.25) | 1.00 (0.90–1.12) |
| 1 gap vs 0 gaps | Stroke | 1.95 (1.54–2.45) | 1.46 (1.28–1.67) | 1.14 (0.91–1.44) |
| 2 gaps vs 0 gaps | CVD | 1.67 (1.43–1.96) | 1.30 (1.17–1.44) | 1.20 (1.06–1.36) |
| 2 gaps vs 0 gaps | Heart disease | 1.30 (1.08–1.57) | 1.25 (1.12–1.40) | 1.13 (0.99–1.29) |
| 2 gaps vs 0 gaps | Stroke | 2.76 (2.10–3.62) | 1.53 (1.26–1.87) | 1.38 (1.06–1.81) |
| 3 gaps vs 0 gaps | CVD | 2.49 (1.86–3.33) | 1.93 (1.54–2.40) | 1.23 (0.90–1.67) |
| 3 gaps vs 0 gaps | Heart disease | 1.96 (1.39–2.78) | 1.80 (1.41–2.28) | 1.13 (0.82–1.56) |
| 3 gaps vs 0 gaps | Stroke | 4.61 (2.98–7.16) | 2.49 (1.64–3.76) | 1.57 (0.85–2.90) |

Exact gap count is the total number of U, RU, or TA states across the 3 targets. Model covariates and variance estimators are described in Section 2.3 and Table S1C.

Gap count was also positively associated with stroke, with HRs per additional gap of 1.63 (1.46–1.83), 1.30 (1.20–1.41), and 1.18 (1.03–1.33) in CHARLS, HRS, and ELSA, respectively. For heart disease, the corresponding HRs were 1.18 (1.08–1.28), 1.15 (1.09–1.21), and 1.06 (0.99–1.13).

### 3.3 Overall Care Phenotypes and Stage Differences Among Single Gaps

Compared with overall G0, single-gap TA and multiple-gap M were associated with higher CVD risk in each cohort. Single-gap U had CVD HRs close to 1, while RU was associated with higher CVD risk in CHARLS and HRS. U was also associated with higher stroke risk in CHARLS and HRS (Figure 3B and Table 2B).

**Figure 3.**
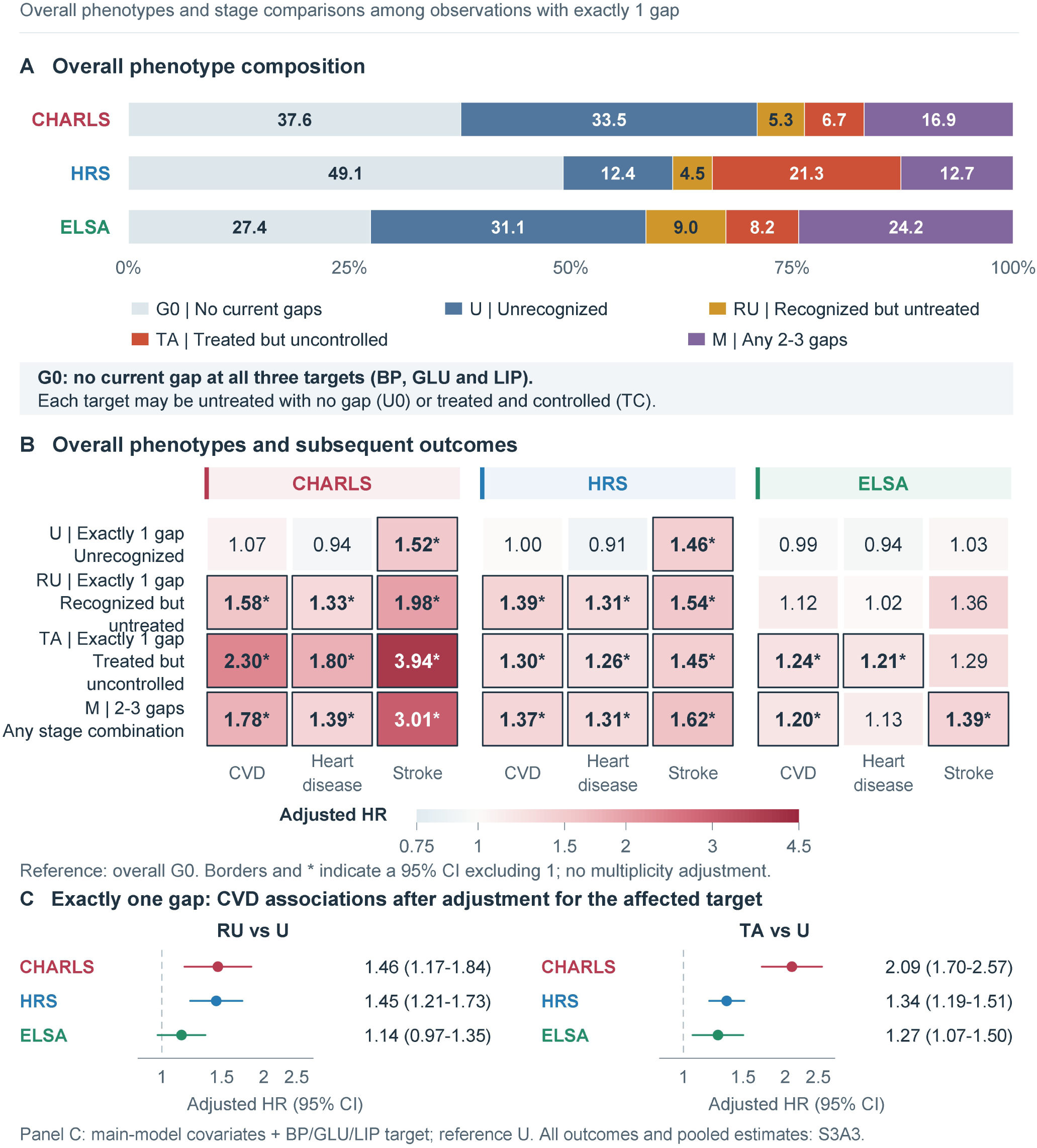
Overall care phenotypes and single-gap stage comparisons. A, Unweighted phenotype distribution in the CVD sample. B, Adjusted HRs for U, RU, TA, and M versus overall G0; borders and asterisks indicate pointwise 95% CIs excluding 1, without adjustment for multiple comparisons. U, RU, and TA denote a single unrecognized, recognized but untreated, or treated but uncontrolled care gap, respectively. M includes all stage combinations with 2–3 gaps. G0 requires no current gap in any of the 3 targets; each target may be U0 or TC. C, Associations of RU and TA with CVD among observations with exactly 1 gap, using U as the reference and additionally adjusting for the affected target. Full results are provided in Table 2B and Tables S3A1–S3A3.

**Table 2B.** Adjusted Associations of Overall Care Phenotypes With Cardiovascular Outcomes.

| **Phenotype (vs G0)** | **Outcome** | **CHARLS HR (95% CI)** | **HRS HR (95% CI)** | **ELSA HR (95% CI)** |
|---|---|---|---|---|
| Single unrecognized gap (U) | CVD | 1.07 (0.93–1.23) | 1.00 (0.90–1.11) | 0.99 (0.88–1.11) |
| Single unrecognized gap (U) | Heart disease | 0.94 (0.80–1.10) | 0.91 (0.80–1.02) | 0.94 (0.83–1.06) |
| Single unrecognized gap (U) | Stroke | 1.52 (1.18–1.97) | 1.46 (1.20–1.76) | 1.03 (0.80–1.34) |
| Single recognized but untreated gap (RU) | CVD | 1.58 (1.26–1.97) | 1.39 (1.19–1.63) | 1.12 (0.95–1.33) |
| Single recognized but untreated gap (RU) | Heart disease | 1.33 (1.02–1.73) | 1.31 (1.10–1.55) | 1.02 (0.85–1.22) |
| Single recognized but untreated gap (RU) | Stroke | 1.98 (1.32–2.97) | 1.54 (1.14–2.08) | 1.36 (0.96–1.93) |
| Single treated but uncontrolled gap (TA) | CVD | 2.30 (1.92–2.76) | 1.30 (1.20–1.40) | 1.24 (1.06–1.46) |
| Single treated but uncontrolled gap (TA) | Heart disease | 1.80 (1.45–2.24) | 1.26 (1.16–1.38) | 1.21 (1.03–1.44) |
| Single treated but uncontrolled gap (TA) | Stroke | 3.94 (2.92–5.31) | 1.45 (1.25–1.69) | 1.29 (0.90–1.84) |
| Multiple care gaps (M) | CVD | 1.78 (1.53–2.07) | 1.37 (1.24–1.51) | 1.20 (1.06–1.36) |
| Multiple care gaps (M) | Heart disease | 1.39 (1.16–1.66) | 1.31 (1.18–1.46) | 1.13 (0.99–1.29) |
| Multiple care gaps (M) | Stroke | 3.01 (2.31–3.92) | 1.62 (1.34–1.96) | 1.39 (1.06–1.82) |

The reference is overall G0. Overall U, RU, and TA are restricted to exactly 1 gap; M includes same-stage and mixed-stage combinations with multiple gaps. Direct single-gap comparisons and additional target adjustment are reported in Tables S3A2–S3A3; multiple-gap comparisons are reported in Tables S3C1–S3C2.

With exactly 1 gap, CVD risk was higher for TA than for U even after adjustment for the affected target. HRs were 2.09 (1.70–2.57) in CHARLS, 1.34 (1.19–1.51) in HRS, and 1.27 (1.07–1.50) in ELSA (Figure 3C). All 3 cohort-specific HRs exceeded 1, although their magnitudes differed substantially. The pooled HR was 1.52 (0.77–2.97), with I²=90.3%. A similar pattern was observed for heart disease. Comparisons involving RU and those for stroke varied across cohorts (Figure S2 and Tables S3A2–S3A3).

Distinguishing U, RU, and TA within the single-gap category improved model fit for CVD and heart disease beyond exact gap count in all 3 cohorts. The joint Wald tests remained significant after Holm correction. For stroke, this stage distinction was supported only in CHARLS (Figure 4 and Table 3). These model comparisons used the full analysis sample for each outcome. Figure 4 and Tables S3B1–S3B2 report gains in model fit and apparent ΔC-index values.

**Table 3.** Information From Single-Gap Care Stage Beyond Gap Count.

| Cohort | Outcome | LR χ² gain<br>Mean (95% interval) | Partial R² gain<br>Mean (95% interval) | Wald χ²<br>(df) | Holm P |
|---|---|---|---|---|---|
| CHARLS | CVD | 68.90 (38.87–110.73) | 0.0087 (0.0049–0.0139) | 72.04 (2) | <0.001 |
| CHARLS | Heart disease | 35.84 (14.29–61.99) | 0.0045 (0.0018–0.0078) | 36.97 (2) | <0.001 |
| CHARLS | Stroke | 42.01 (17.32–73.55) | 0.0053 (0.0022–0.0093) | 44.84 (2) | <0.001 |
| HRS | CVD | 30.23 (12.01–53.99) | 0.0015 (0.0006–0.0026) | 24.01 (2) | <0.001 |
| HRS | Heart disease | 37.39 (15.87–63.07) | 0.0018 (0.0008–0.0031) | 27.85 (2) | <0.001 |
| HRS | Stroke | 2.28 (0.06–8.12) | 0.0001 (0.0000–0.0004) | 0.16 (2) | 0.925 |
| ELSA | CVD | 14.90 (2.36–34.45) | 0.0012 (0.0002–0.0029) | 8.65 (2) | 0.037 |
| ELSA | Heart disease | 14.40 (1.99–31.70) | 0.0012 (0.0002–0.0026) | 8.80 (2) | 0.037 |
| ELSA | Stroke | 6.99 (0.29–21.54) | 0.0006 (0.0000–0.0018) | 3.38 (2) | 0.184 |

All comparisons are M3−M1. LR χ² and partial R² are means across 500 bootstrap resamples with pointwise 95% percentile intervals. Wald tests use the variance estimator of each cohort's primary model. Holm correction treats the 3 outcomes within each cohort and comparison as a family. Other model comparisons and apparent ΔC-index values are reported in Tables S3B1–S3B2.

**Figure 4.**
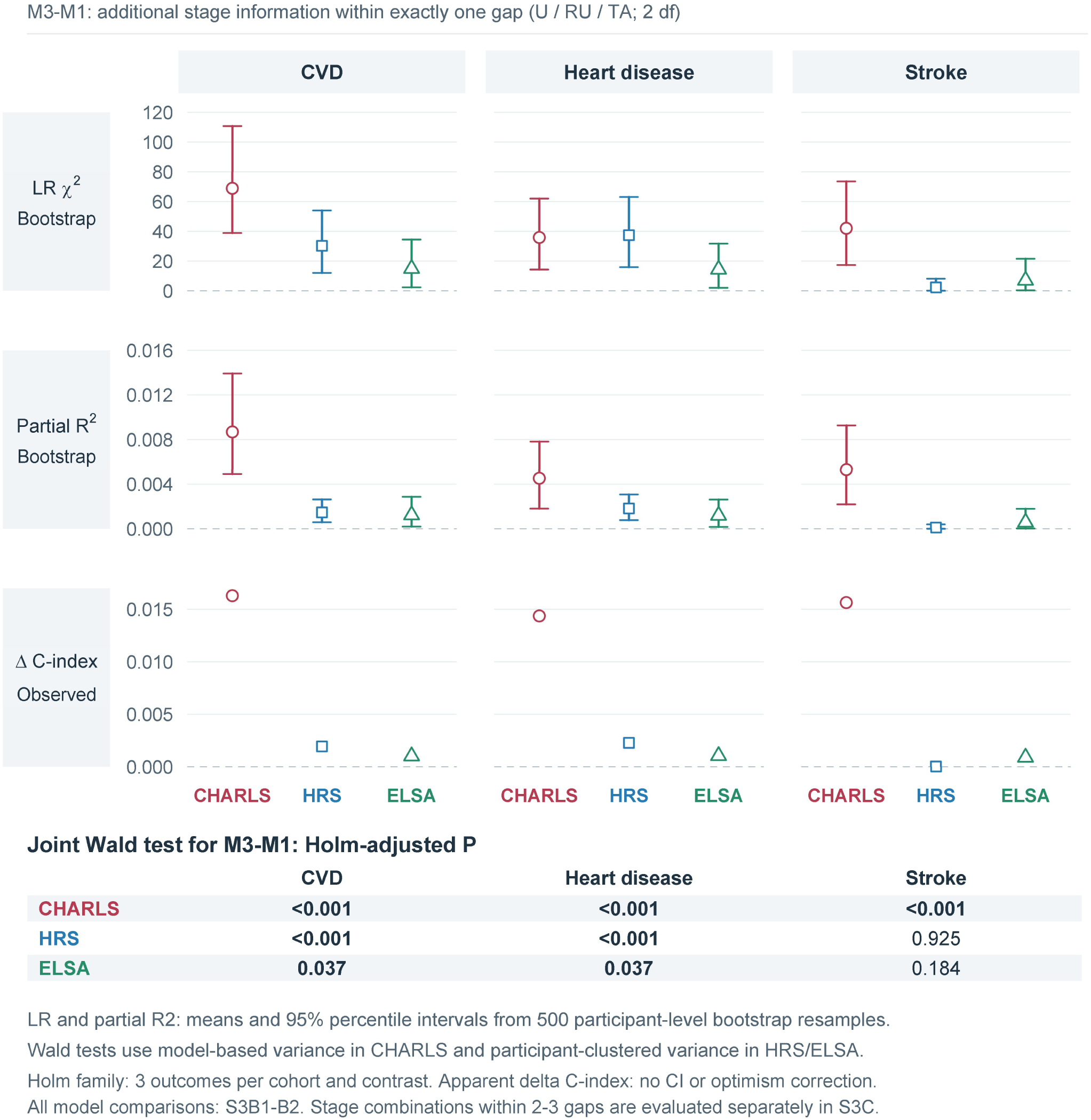
Information from single-gap care stage beyond gap count. The figure shows M3−M1: M1 includes covariates and exact gap count; M3 also includes overall phenotype, adding 2 degrees of freedom for stage among observations with exactly 1 gap. The first 2 rows show mean gains in LR χ² and partial R² across 500 participant-level bootstrap resamples with pointwise 95% percentile intervals. The third row shows the apparent ΔC-index in the same sample, without a CI or correction for optimism. Joint Wald tests with Holm-corrected P values are shown below, treating the 3 outcomes in each cohort as a family. CHARLS uses model-based variance, and HRS and ELSA use participant-clustered robust covariance. Full comparisons are provided in Tables S3B1–S3B2.

Exact gap count also provided information beyond stage classification. In CHARLS and HRS, distinguishing 2 from 3 gaps within M improved the models for CVD, heart disease, and stroke by joint Wald tests, with all comparisons remaining significant after Holm correction. The corresponding comparisons in ELSA were not significant (Table S3B2).

### 3.4 Stage Characteristics Among Observations With Multiple Gaps

At a fixed gap count, outcomes did not differ consistently between same-stage and mixed-stage combinations. For 2 gaps, the pooled CVD HR for mixed-stage versus same-stage combinations was 1.06 (0.73–1.53). For 3 gaps, it was 0.91 (0.66–1.24). Findings were similar for heart disease and stroke (Figure S3 and Tables S3C1–S3C2).

Among observations with exactly 2 gaps, combinations containing TA were associated with higher CVD risk in each cohort. After adjustment for target composition, HRs for combinations containing TA versus those without TA were 1.63 (1.29–2.07) in CHARLS, 1.69 (1.27–2.24) in HRS, and 1.25 (1.05–1.49) in ELSA. The pooled HRs (pointwise 95% CIs) were 1.48 (0.97–2.24) for CVD and 1.47 (1.12–1.92) for heart disease. The Holm-corrected P value for heart disease was 0.077. Results for CVD and heart disease were broadly similar when observations with 2–3 gaps were combined and exact gap count was adjusted for. Stroke comparisons and comparisons restricted to exactly 3 gaps varied across cohorts (Tables S3C1–S3C2).

### 3.5 Target-Specific Care Stages and Cardiovascular Outcomes

Unrecognized abnormalities had different associations with subsequent outcomes across targets and cohorts. In CHARLS, unrecognized glycemic gaps were associated with CVD and heart disease relative to U0, with HRs of 1.62 (1.25–2.10) and 1.64 (1.21–2.22), respectively. Corresponding estimates in HRS and ELSA were close to 1. Unrecognized blood pressure gaps were associated with higher stroke risk in CHARLS and HRS, with HRs of 1.80 (1.36–2.37) and 1.77 (1.41–2.21), respectively (Figure 5 and Table S5A3).

**Figure 5.**
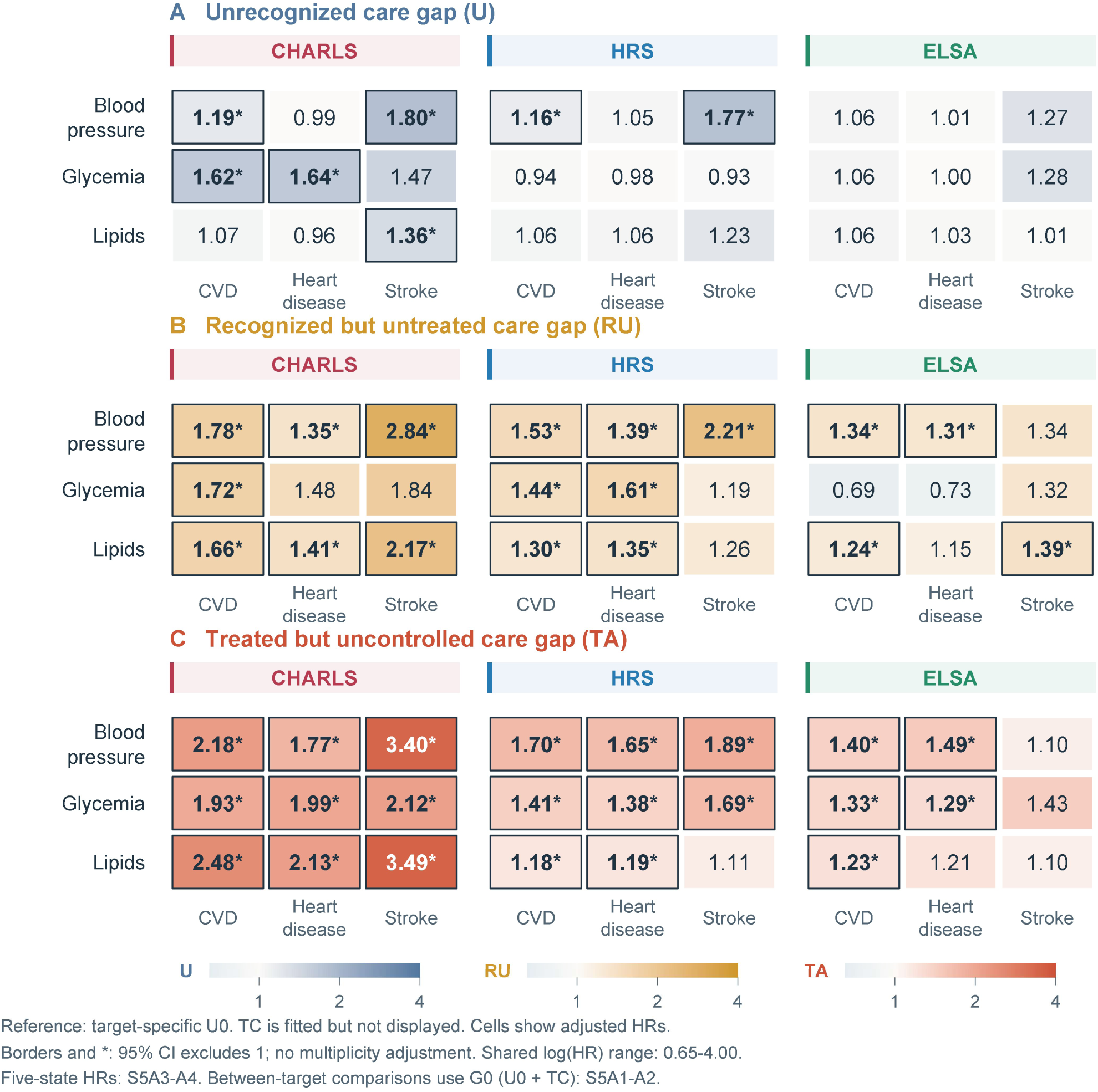
Associations of blood pressure, glycemic, and lipid care stages with cardiovascular outcomes. A, B, and C show adjusted HRs for unrecognized (U), recognized but untreated (RU), and treated but uncontrolled (TA) care gaps, respectively. The reference is untreated with no current gap (U0); treated and currently controlled (TC) is included in the full 5-state models. Colors map to log(HR); borders and asterisks indicate pointwise 95% CIs excluding 1, without adjustment for multiple comparisons. CHARLS uses model-based variance, and HRS and ELSA account for participant clustering. Full results are provided in Tables S5A3–S5A4. Formal comparisons across targets are reported in Tables S5A1–S5A2, using target-specific G0 as the reference.

Recognized but untreated blood pressure gaps were associated with higher CVD risk in all 3 cohorts. HRs were 1.78 (1.41–2.24) in CHARLS, 1.53 (1.25–1.88) in HRS, and 1.34 (1.13–1.60) in ELSA. TA was associated with higher CVD risk for each target in every cohort. These estimates and Figure 5 come from 5-state models using U0 as the reference. Full results are reported in Tables S5A3–S5A4.

Direct comparisons among targets used target-specific G0, which combines U0 and TC, as the common reference definition rather than U0 alone. In CHARLS, the CVD association for unrecognized glycemic gaps was stronger than those for unrecognized blood pressure and lipid gaps. RHRs were 0.65 (0.47–0.88) for blood pressure versus glycemia and 1.57 (1.19–2.08) for glycemia versus lipids. The overall heart disease test also identified differences among targets.

In HRS, associations differed among targets at the TA stage. For CVD, RHRs were 1.21 (1.07–1.36) for blood pressure versus lipids and 1.23 (1.08–1.41) for glycemia versus lipids. Blood pressure and glycemic associations were also stronger than lipid associations for heart disease, while the glycemic association was stronger than the lipid association for stroke. The overall tests and these pairwise comparisons remained significant after Holm correction. Corresponding tests in ELSA did not identify within-stage differences among targets (Figure S5A and Tables S5A1–S5A2).

CVD HRs for TA were closer to 1 when TC rather than U0 was used as the reference. Pooled TA versus TC HRs were 1.11 (0.98–1.26) for blood pressure, 1.10 (0.92–1.33) for glycemia, and 1.03 (0.98–1.08) for lipids. For blood pressure in HRS, the TA HR was 1.70 (1.53–1.89) relative to U0 and 1.14 (1.05–1.24) relative to TC. The proportion of treated and controlled observations within each target’s G0 also differed across cohorts. Figures S4A–S4B and Tables S4A–S4B show the composition of G0 and the estimates under both reference groups.

Some overall phenotype associations remained after individual targets were omitted. With blood pressure omitted, the pooled HR for M and stroke was 1.89 (1.08–3.30). With lipids omitted, RU remained associated with higher CVD risk, with a pooled HR of 1.33 (1.08–1.64). Associations with heart disease and stroke followed the same direction (Figure S5B and Table S5B).

### 3.6 Additional Analyses

Results for gap count, single-gap TA, and M were broadly similar with common minimal adjustment, spline modeling of age and BMI, and fixed-effect versus random-effects meta-analysis. Omitting 1 cohort at a time changed the magnitude of some pooled estimates (Figure S6 and Tables S6A1–S6B4).

In CHARLS, CVD associations for gap count, single-gap TA, and M remained positive after requiring a 2011 visit or classifiable gaps in that year. All these analyses required confirmed absence of CVD in 2015. Estimates for U and some stage comparisons changed more substantially (Table S7G2). In HRS, excluding observations with a current gap and missing treatment responses had little effect on overall phenotype HRs (Table S7H1). Requiring complete urban–rural residence data reduced CVD observations from 20,531 to 19,905, again with little change in overall phenotype HRs (Table S7H2).

Direct comparisons of stroke and heart disease identified stronger stroke associations for U, TA, and M in CHARLS and for U in HRS. These comparisons remained significant after within-cohort Holm correction. The RHR for TA in CHARLS was 2.15 (1.48–3.13). When estimates were pooled across the 3 cohorts, none of the 4 phenotype comparisons remained significant after Holm correction (Table S5C).

Several associations changed over follow-up. In the HRS lipid 5-state models, exposure-level proportional hazards tests indicated time variation for CVD and heart disease, mainly for U versus U0. The CVD HR for this comparison changed from 0.49 (0.27–0.87) at year 1 to 1.16 (0.97–1.40) at year 5, with a similar pattern for heart disease. The association of 3 versus 0 gaps with stroke in HRS also attenuated over time. The main tables report HRs that summarize associations over the full follow-up period. Time-specific estimates and complete tests are provided in Tables S6B5–S6B9.

## 4 Discussion

In 3 cohorts of middle-aged and older adults, a greater number of unresolved blood pressure, glycemic, and lipid care gaps was generally associated with higher subsequent CVD risk. Among observations with only 1 unresolved target, TA was associated with higher CVD risk than U in each cohort, even after adjustment for the affected target. Within the same care stage, associations also differed by target in some cohorts. Unrecognized glycemic gaps had stronger CVD associations than unrecognized blood pressure or lipid gaps in CHARLS, while blood pressure and glycemic gaps had stronger associations than lipid gaps at the TA stage in HRS. For multiple gaps, grouping combinations as same-stage or mixed-stage did not consistently distinguish outcomes. With exactly 2 gaps, however, combinations containing TA were associated with higher CVD risk in each cohort.

Previous studies have largely considered the burden of coexisting cardiometabolic risk factors^15–19^ or the stages of recognition, treatment, and control.^9–13^ Work in hypertension has also examined the prognostic associations of these stages.^14^ We addressed both questions by defining care gaps as current unresolved problems. An unrecognized abnormality meeting the cohort-specific gap criteria counted as a gap, while a target meeting the corresponding control criteria did not. We then tested whether stage added information beyond exact gap count and whether count remained informative after stage classification. Together with the comparisons of stage combinations, these findings support a simpler description of care needs based on count, stage, and affected target.

Single gaps were the most common pattern in all 3 cohorts, accounting for approximately two-thirds to three-quarters of observations with any gap. With the count held constant, TA remained associated with higher CVD risk than U after the affected target was taken into account. Care stage reflects prior recognition and treatment as well as the problem that remains unresolved.

Exact gap count also remained informative after care stage was considered. In CHARLS and HRS, outcomes differed between 2 and 3 gaps after accounting for stage, whereas the corresponding comparisons in ELSA were not significant. The proposed count–stage sequence organizes assessment around the number of unresolved targets and then their care stages. The prognostic contribution of each dimension varies across cohorts.

For multiple gaps, outcomes did not differ consistently between same-stage and mixed-stage combinations at a fixed count. The same-stage category includes both 2 unrecognized abnormalities and 2 treated but uncontrolled abnormalities. A mixed-stage combination might include an unrecognized lipid abnormality alongside blood pressure that remains uncontrolled during treatment, calling for diagnostic confirmation of the lipid finding and reassessment of blood pressure control. Among observations with exactly 2 gaps, those containing TA had higher CVD risk in each cohort, although the pooled estimate remained imprecise. These findings support detailed stage descriptions for single gaps. A more parsimonious description of multiple gaps could retain exact count and affected targets while recording whether any target remains uncontrolled during treatment.

Unrecognized lipid gaps were present in approximately two-fifths of CHARLS participants and ELSA landmark observations. In CHARLS, however, unrecognized glycemic gaps had a stronger CVD association than unrecognized lipid gaps. The CHARLS finding is consistent with previous reports of limited dyslipidemia awareness in China.^25–26^ Gap frequency describes how common an unresolved problem was in the analyzed sample. States with stronger outcome associations may warrant closer clinical review, while care stage identifies whether that review should focus on diagnosis, treatment initiation, or control during treatment.

Unrecognized blood pressure gaps were associated with higher stroke risk in CHARLS and HRS. Recognized but untreated blood pressure gaps were associated with higher CVD risk in all 3 cohorts. Previous research in chronic coronary syndromes has also linked blood pressure control to cardiovascular risk.^27^ For a recognized but untreated gap, the existing diagnosis provides a starting point for reviewing why treatment has not begun and whether it is indicated.^5^ For an unrecognized gap, follow-up could focus on reassessing the abnormal measurement and confirming the diagnosis.

Under each cohort’s prespecified definition, TA accounted for the largest proportion of glycemic gaps, making control during treatment the most common unresolved glycemic care problem. Unrecognized glycemic gaps were also associated with higher CVD and heart disease risk in CHARLS. Diagnostic assessment remains relevant alongside evaluation of ongoing treatment and control.^6–7^

In HRS, associations differed by target even at the same care stage. At the TA stage, CVD associations relative to each target’s G0 were stronger for blood pressure and glycemia than for lipids. A similar pattern was observed for heart disease, while the glycemic association was stronger than the lipid association for stroke. To examine whether any one target accounted for the overall phenotype associations, we omitted each target in turn. Several associations persisted. After blood pressure was omitted, multiple gaps involving glycemia and lipids remained associated with stroke. After lipids were omitted, RU remained associated with CVD. These findings suggest that the overall associations were not driven entirely by a single target.

The choice of reference group helps explain the higher HRs for TA. Its CVD HRs were higher relative to U0 and closer to 1 relative to TC. TA and TC both include a history of treatment, whereas U0 describes an untreated state with no current gap. The TA comparisons therefore describe prognosis in people with different disease and treatment histories, alongside differences in current control. Target-specific G0 combines U0 and TC, and overall G0 permits either state at each of the 3 targets. G0 identifies the absence of an unresolved gap at that assessment, while allowing for differences in underlying risk and follow-up needs.

We harmonized care states across cohorts while allowing the underlying measurements and thresholds to differ. HRS lipid classification used total cholesterol and HDL-C, while CHARLS and ELSA also used LDL-C and triglycerides. Glycemic indicators, units, and thresholds also differed. In each cohort, the available measurements, diagnoses, and treatment information were used to determine whether a gap was present and whether it concerned recognition, treatment, or control. This preserved a common description of care needs across cohorts with different biomarker panels.

Across the 3 cohorts, gap count was positively associated with CVD, and care stage provided additional information among observations with a single gap. The strength of these associations varied across cohorts. The pooled TA versus U comparison had high heterogeneity, and the target distinctions were most apparent for unrecognized glycemic gaps in CHARLS and TA blood pressure and glycemic gaps in HRS. These findings support applying the same conceptual care framework across populations while recognizing that association estimates and target-specific patterns may remain population-specific. The relative associations with stroke and heart disease also varied by cohort, and pooled comparisons were not significant after Holm correction.

Some associations varied over follow-up. In HRS, the lipid U versus U0 association changed over time, while the association of 3 versus 0 gaps with stroke attenuated. A state recorded at the landmark may therefore have different associations with near-term and longer-term outcomes. It describes the unresolved problems at that visit, after which diagnosis, treatment, and control may change. Repeated measurements and prescription records could distinguish persistent gaps from newly recognized or resolved problems. Future longitudinal studies could relate changes in gap count, duration of control, and transitions between stages to subsequent outcomes, and examine how reassessment intervals affect the detection of these changes.

### Strengths and Limitations

This study combined current measurements with diagnosis and treatment histories to define care gaps and examined their prospective associations in 3 independent cohorts. It compared stage at a fixed count, accounted for the affected target in single-gap comparisons, and directly compared targets within the same stage. Comparisons with both untreated and treated reference states helped interpret the TA associations. The main directions of association were similar across covariate sets, functional forms for age and BMI, and pooling methods. Omitting individual cohorts showed how each influenced the pooled estimates. Together, the distributions and outcome analyses show which care gaps were common and which were associated with greater subsequent risk.

Care states were defined from measurements, reported diagnoses, and medication information available at the assessment. Limited information on disease duration, dose, and adherence may have left residual confounding and some misclassification. Outcomes were reported in subsequent surveys, and the survey at first report approximated event time. The items comprising heart disease differed across cohorts, which may partly explain differences in association magnitude. Competing mortality was not modeled uniformly across the 3 cohorts.

Selection into the complete-case samples differed across cohorts. In ELSA, included observations were younger than excluded observations, with a standardized mean difference in age of −0.373 (Table S7B). The analyses estimated associations between care states and subsequent outcomes within each cohort, and unweighted proportions describe the observations included in those analyses. The populations represented by these samples reflect the available measurements, classification thresholds, and selection process.

## 5 Conclusions

Gap count and care stage provided complementary prognostic information across the 3 cohorts. Within the same stage, associations also varied by target in some cohorts. Care stage added information for single gaps, while a compact description of multiple gaps could retain exact count, affected targets, and the presence of TA. The count–stage framework offers a structured way to identify which cardiometabolic problems remain unresolved and where further assessment is needed.

## ARTICLE INFORMATION

### Ethics Approval and Informed Consent

Original HRS data collection was approved by the University of Michigan Institutional Review Board. ELSA waves and relevant components received approval from the appropriate research ethics committees, and CHARLS was approved by the Peking University ethics review committee. All 3 cohorts obtained informed consent before participation. This study used deidentified secondary data obtained under the access and use requirements of each cohort and involved no recontact with participants.

### Data and Code Availability

CHARLS, HRS, and ELSA data can be requested or accessed through the respective study websites under their data access procedures: CHARLS (https://charls.pku.edu.cn/en/), HRS (https://hrs.isr.umich.edu/), and ELSA (https://www.elsa-project.ac.uk/). We do not redistribute person-level data governed by the original cohort data-use agreements. Harmonization rules, analysis scripts, and nonidentifying summary results are available from the corresponding author upon reasonable request, subject to the original cohort agreements.

### Author Contributions

Yunxi Yang: Conceptualization, Methodology, Investigation, Data curation, Visualization, and Writing—original draft. Jia Lv: Investigation, Data curation, Validation, and Writing—review and editing. Xinyi Tang: Investigation, Data curation, and Validation. Xinghua Zheng: Investigation, Data curation, and Validation. Xing Liu: Conceptualization, Supervision, and Writing—review and editing. All authors read and approved the final manuscript.

## Acknowledgments

The authors thank the CHARLS, HRS, and ELSA research teams and all participants for their contributions to data collection, management, and sharing.

## Supplemental Material

Supplemental Methods

Supplemental Tables

Supplemental Figures

## Nonstandard Abbreviations and Acronyms

CHARLS: China Health and Retirement Longitudinal Study
ELSA: English Longitudinal Study of Ageing
G0: no current gap at the target level; no current gaps across all 3 targets at the overall level
HRS: Health and Retirement Study
M: multiple care gaps (2–3 current gaps)
RHR: ratio of hazard ratios
RU: recognized but untreated care gap
TA: treated but uncontrolled care gap
TC: treated and currently controlled
U: unrecognized care gap
U0: untreated with no current gap

